# Strong Correlation Between Prevalence of Severe Vitamin D Deficiency and Population Mortality Rate from COVID-19 in Europe

**DOI:** 10.1101/2020.06.24.20138644

**Authors:** Isaac Z. Pugach, Sofya Pugach

## Abstract

**Background:** SARS-CoV-2 virus causes a very wide range of COVID-19 disease severity in humans: from completely asymptomatic to fatal, and the reasons behind it are often not understood. There is some data that Vitamin D may have protective effect, so authors decided to analyze European country-wide data to determine if Vitamin D levels are associated with COVID-19 population death rate.

**Methods:** To retrieve the Vitamin D levels data, authors analyzed the Vitamin D European population data compiled by 2019 ECTS Statement on Vitamin D Status published in the European Journal of Endocrinology. For the data set to used for analysis, only recently published data, that included general adult population of both genders ages 40-65 or wider, and must have included the prevalence of Vitamin D deficiency.

**Results:** There were 10 countries data sets that fit the criteria and were analyzed. Severe Vitamin D deficiency was defined as 25(OH)D less than 25 nmol/L (10 ng/dL). Pearson correlation analysis between death rate per million from COVID-19 and prevalence of severe Vitamin D deficiency shows a strong correlation with r = 0.76, p = 0.01, indicating significant correlation. Correlation remained significant, even after adjusting for age structure of the population. Additionally, over time, correlation strengthened, and r coefficient asymptoticaly increased.

**Conclusions:** Authors recommend universal screening for Vitamin D deficiency, and further investigation of Vitamin D supplementation in randomized control studies, which may lead to possible treatment or prevention of COVID-19.

## Introduction

SARS-CoV-2 virus causes a very wide range of disease severity in humans: from completely asymptomatic to fatal, and the reasons behind it are often not understood. There is some data that Vitamin D may have protective effect (1, 2), so authors decided to analyze European country-wide data to determine if Vitamin D deficiency is associated with COVID-19 population death rate.

## Methods

To retrieve the Vitamin D levels data, authors analyzed European countries Vitamin D population data compiled by 2019 ECTS Statement on Vitamin D Status published in the European Journal of Endocrinology (3) For the data set to used for analysis, it must have been published in the last 10 years, must have included general adult population of both genders ages 40-65 or wider, and must have had breakdown on the prevalence of Vitamin D deficiency (not just the mean value).

To determine the number of deaths from COVID-19, authors retrieved the country data from the John Hopkins University of Medicine Coronavirus Resource Center (4). To determine the 2020 demographics data, authors retrieved the country data from CIA World Factbook (5). Four data retrievals were performed between May 11, 2020 and June 14, 2020.

Pearson correlation analysis was performed on the resulting data. Additionally, regression analysis was performed to adjust the death rate for age structure of each country. LibreOffice Calc software was used for statistical analysis.

## Results

**Table 1.** **Vitamin 25(OH) D levels in general adult population and mortality rates per million from COVID-19 in European countries, and their correlation statistics.**

| Country | Prevalence (%) of<br>Vit. D < 25 nmol/L | Prevalence (%) of<br>Vit. D < 50 nmol/L | Mean (nmol/L)<br>Vitamin D | Population | COVID-19<br>Deaths | COVID-19<br>Deaths/million |
| --- | --- | --- | --- | --- | --- | --- |
| UK | 15.4 | 56.4 | 47.4 | 65,761,117 | 41,747 | 635 |
| France | 6.3 | 34.6 | 60 | 67,848,156 | 29,401 | 433 |
| Belgium | 7.3 | 51.1 | 49.3 | 11,720,176 | 9,655 | 824 |
| Germany | 4.2 | 54.5 | 50.1 | 80,159,662 | 8,801 | 110 |
| Netherlands | 4.9 | 33.6 | 59.5 | 17,280,397 | 6,078 | 352 |
| Ireland | 6 | 45 | 56.4 | 5,176,569 | 1,705 | 329 |
| Denmark | 0 | 23.6 | 65 | 5,869,410 | 597 | 102 |
| Finland | 0.2 | 6.6 | 67.7 | 5,571,665 | 326 | 59 |
| Norway | 0.3 | 18.6 | 65 | 5,467,439 | 242 | 44 |
| Estonia | 4.5 | 51 | 51.6 | 1,228,624 | 69 | 56 |
| N= 10 |  |  |  |  |  |  |
| **Correlation coefficient** | **0.76** | **0.52** | **-0.57** |  |  |  |
| **T-statistic** | **3.33** | **1.72** | **-1.98** |  |  |  |
| **P-value** | **0.01** | **0.12** | **0.08** |  |  |  |

Data retrieved June 14, 2020

Pearson correlation analysis shows that severe Vitamin D deficiency (defined as serum Vitamin 25(OH) D less than 25 nmol/L) is strongly and significantly correlated with COVID-19 deaths per million of population with r = 0.76, p = 0.01 (95% confidence interval 0.25 − 0.81). After adjusting for population age structure of the countries, the correlation remained strong and significant, with partial correlation coefficient r = 0.76, and p = 0.03.

Prevalence of mild Vitamin D deficiency (defined as serum Vitamin 25(OH) D less than 50 nmol/L) is moderately correlated with COVID-19 deaths per million, but this correlation does not reach statistical significance, with r = 0.52, p = 0.12. Vitamin D levels are inversely correlated with the COVID-19 deaths per million, with r = −0.57, p = 0.08.

Over a course of about a month, the authors retrieved the COVID-19 deaths data, and performed the correlation analysis. As demonstrated by Graph 2, correlation coefficient r is asymptotically increasing over time, with the correlation becoming stronger.

**Graph 1.**
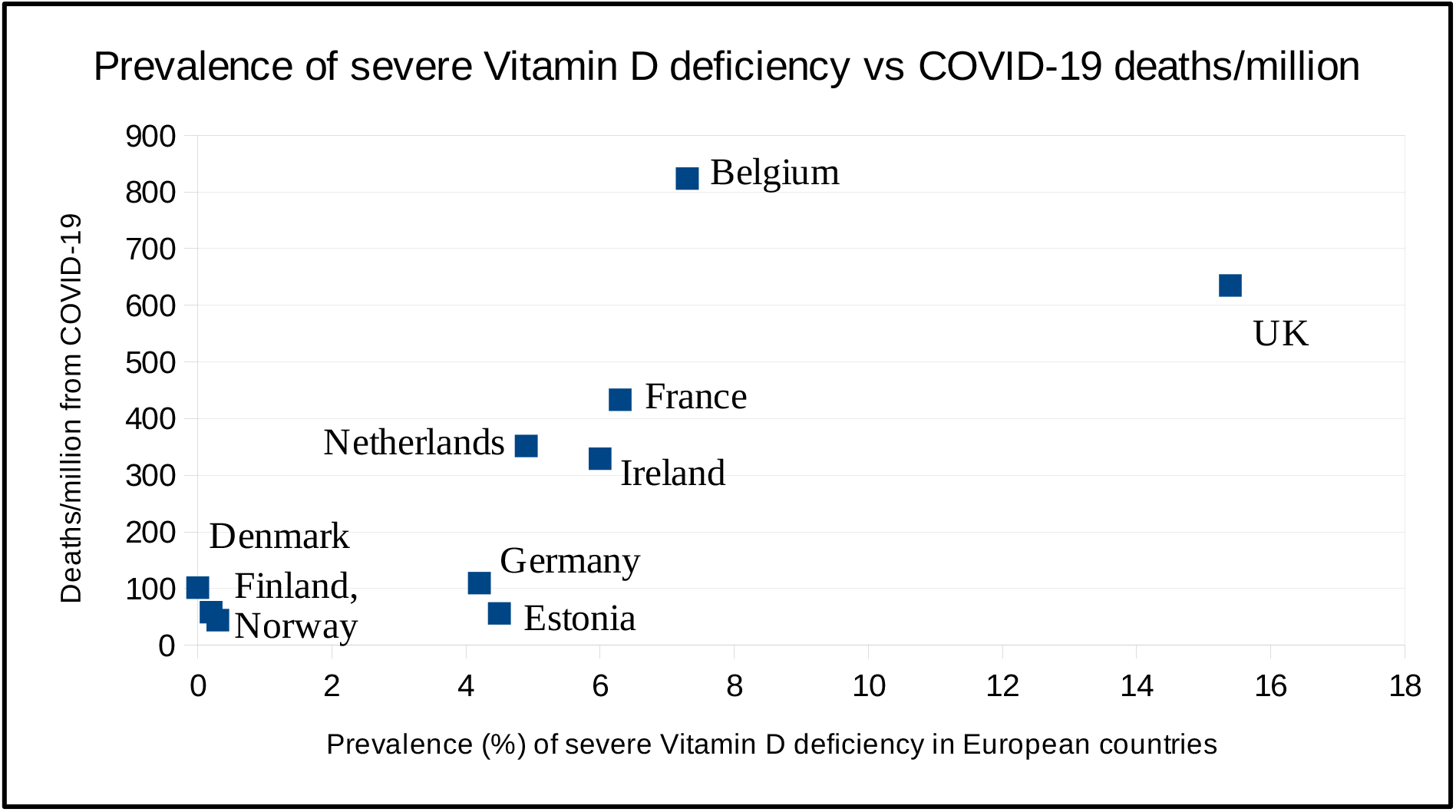
Prevalence of Severe Vitamin D deficiency vs. COVID-19 Deaths per Million in Europe.

**Graph 2.**
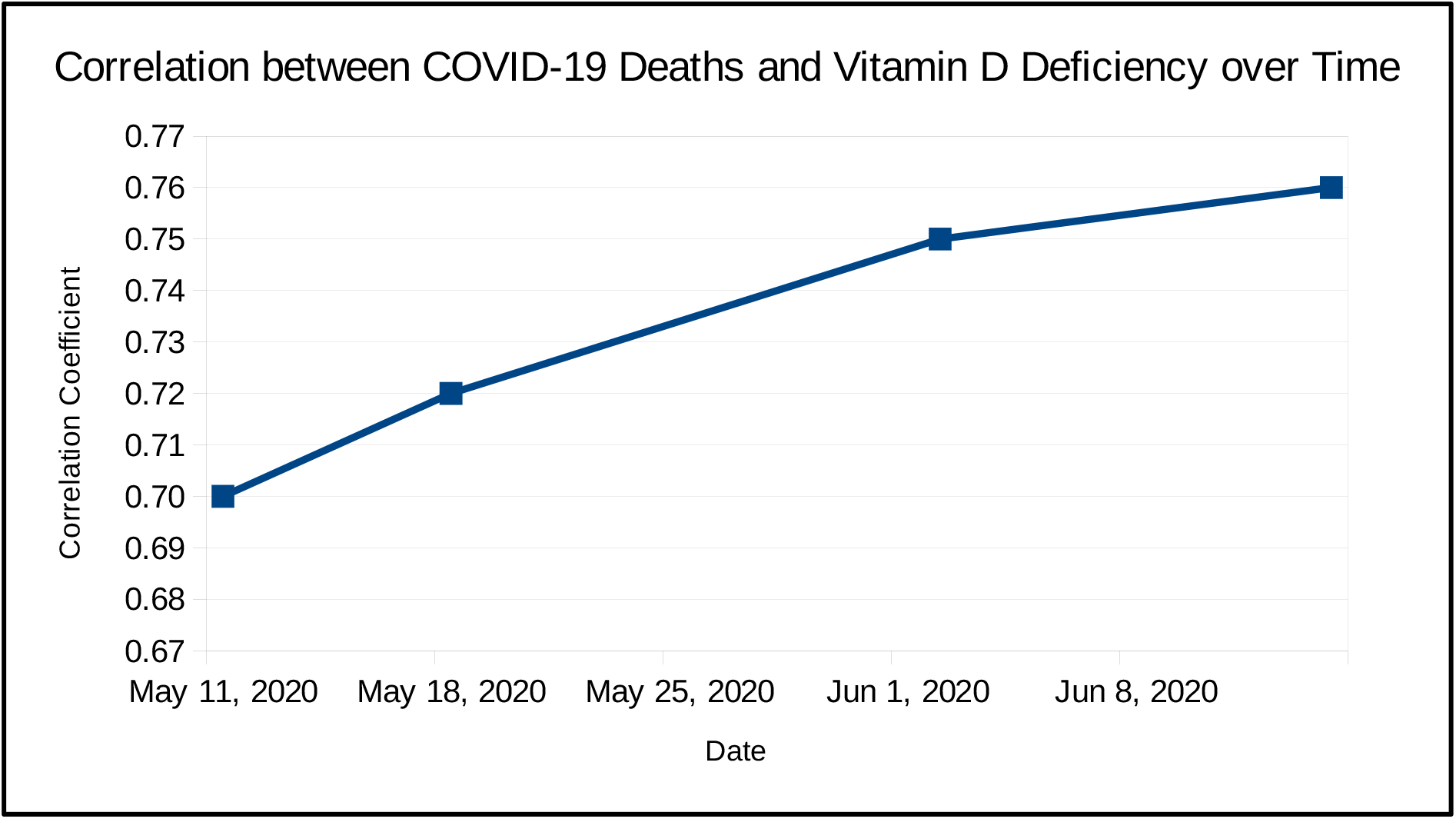
Correlation between COVID-19 Deaths and Vitamin D Deficiency over Time.

## Discussion

The data collected shows a strong and significant correlation between prevalence of severe Vitamin D deficiency and COVID-19 deaths per million in European countries. Moreover, over time, the correlation is increasing asymptotically, making this correlation even less likely to be caused by a random chance.

There is additional data that further supports the correlation between Vitamin D levels and population mortality rate from COVID-19 (6) and shows possible association between low levels of Vitamin D and COVID-19 disease severity (1). Additionally, in the United States, the mortality rate from COVID-19 is higher in blacks and Hispanic Americans than in general population (7,8) and these groups are well known to have lower Vitamin D levels than the general population.

Since the correlation coefficient r = 0.76, it means that about 58% (r2) of death rate from COVID-19 can be explained by prevalence of severe Vitamin D deficiency. This finding by itself does not necessarily mean that low Vitamin D levels increase the death rate, or that that correcting Vitamin D levels would decrease the mortality, because another variable could cause both high prevalence of Vitamin D deficiency and increased deaths from COVID-19. Examples of such variable could be inadequate health care system or prevalence of unknown genetic variation. However, since there is data that Vitamin D deficiency is associated with greater risk and greater severity of various infections (9), a randomized control study that supports the use of Vitamin D to prevent influenza in school children (10), and evidence that supports the use of Vitamin D supplementation for prevention of COVID-19 infections (2), it is very likely that Vitamin D supplements have a large role in prevention and possibly treatment of COVID-19 disease.

## Conclusion

There is a strong correlation between prevalence of severe Vitamin D deficiency and the mortality rate per million from COVID-19 in the European countries, and it is strengthening over time. Authors recommend for physicians to universally screen for Vitamin D deficiency, and recommend further investigation of Vitamin D supplementation in randomized control studies, which may lead to possible treatment or prevention of COVID-19.

*No outside funding was received for this research, and the authors declare that there is no conflicts of interest*.

## Data Availability

The authors have all the data available that was used in the manuscript.

## References

1. Daneshkhah A, Agrawal V, Eshein A et al. The Possible Role of Vitamin D in Suppressing Cytokine Storm and Associated Mortality in COVID-19 Patients. MedRxiv, on line, doi:https://doi.org/10.1101/2020.04.08.20058578. April 2020: 1–9.

2. Grant WB, Lahore H, McDonnell SL et al. Evidence that Vitamin D Supplementation Could Reduce Risk of Influenza and COVID-19 Infections and Deaths. Nutrients. 2020(4): 988–993.

3. Lips P, Cashman KD, Lamberg-Allardt C et al. Current vitamin D status in European and Middle East countries and strategies to prevent vitamin D deficiency: a position statement of the European Calcified Tissue Society. European Journal of Endocrinology, 2019 (4):23–54.

4. John Hopkins University of Medicine Coronavirus Resource Center. Data retrieved on 05/18/2020. https://coronavirus.jhu.edu

5. CIA World Factbook. Data retrieved on 05/18/2020. https://www.cia.gov/library/publications/the-world-factbook

6. Ilie PC, Stefanescu S & Smith, L. The role of vitamin D in the prevention of coronavirus disease 2019 infection and mortality. Aging Clinical and Experimental Research (2020).

7. CDC, Coronavirus disease 2019 (COVID-2019). COVID-19 in Racial and Ethnic Minority groups. https://www.cdc.gov/coronavirus/2019-ncov/need-extra-precautions/racial-ethnic-minorities.html

8. Thebault R, Tran A, & Williams V. The coronavirus is infecting and killing black Americans at an alarmingly high rate. Washington Post. April 7, 2020.

9. Gunville C, Mourani P & Ginde A. The role of Vitamin D in prevention and treatment of infection. Inflamm Allergy Drug Targets. 2013, 12(4):239–245.

10. Urashima M, Segawa T, Okazaki M et al. Randomized trial of vitamin D supplementation to prevent seasonal influenza A in schoolchildren. Am J Clin Nutr. May 2010, 91(5):1255–60.

